# MaternaAI: Enhancing Equitable Maternal Healthcare in Kerala with Fairness-Aware and Explainable Learning Models

**DOI:** 10.64898/2026.08.12.26360340

**Authors:** Ashly Ann Jo, Ebin Deni Raj, Abhijnan Chakraborty

## Abstract

Maternal healthcare prediction systems often inherit biases from imbalanced datasets and socio-economic disparities, undermining their value in equitable healthcare policymaking. We present *MaternaAI*, a fairness-aware and explainable learning framework tailored to enhance maternal healthcare predictions in Kerala, India. The framework focuses on three key indicators: (1) Tetanus Toxoid (TT) booster uptake, (2) immunization coverage, and (3) the percentage of pregnant women completing four or more Antenatal Care (ANC) visits. To address fairness, we propose Adaptive Equity Score Optimization (AESO), a novel, model-agnostic optimization algorithm that dynamically adjusts group equity weights based on real-time disparities. For transparency, *MaternaAI* integrates explainable AI (XAI) techniques, including SHAP, LIME, and feature permutation methods to enable both global and local interpretability. Empirical evaluation using real-world data from Kerala’s Health Management Information System (HMIS) shows that *MaternaAI* improves fairness and predictive accuracy across machine learning and deep learning models, offering actionable, interpretable, and equitable decision support for public health stakeholders.

## 1 INTRODUCTION

Maternal healthcare continues to be a pivotal element of public health strategy, especially in low and middle-income countries like India, where demographic, infrastructural, and socio-economic diversity poses significant challenges to universal health coverage. India’s national maternal health initiatives, under the broader National Health Mission, such as the Janani Suraksha Yojana (JSY) [81], which incentivizes institutional deliveries among economically disadvantaged women, and the Pradhan Mantri Surakshit Matritva Abhiyan (PMSMA) [82], which promotes high-quality antenatal care, have significantly improved maternal health outcomes over the past two decades. Additional programs like LaQshya [47], focused on enhancing labor room quality, and Mission Indradhanush [46], aimed at expanding immunization coverage, further strengthen maternal and child health.

The southern Indian state of Kerala stands out in this national context. It consistently ranks among the best performers in the maternal health indicators, with high rates of institutional deliveries (>99%), low maternal mortality ratio (MMR), and nearly universal immunization coverage [1]. The success can be attributed to a strong Primary Health Centre (PHC) infrastructure, high literacy, decentralized governance, and grassroots community mobilization through programs like Kudumbashree [48]. PHCs in Kerala offer a continuum of maternal healthcare services: from pregnancy registration and routine Antenatal Care (ANC) to nutrition counseling, immunizations, and referrals for high-risk pregnancies [37].

However, this success is not uniformly distributed. Tribal populations in Wayanad, migrant workers in Ernakulam, and coastal communities in Alappuzha face challenges including poor access, language barriers, data under-reporting, and fragmented service continuity [61]. These regional disparities mirror national trends, where marginalized populations are disproportionately impacted by gaps in healthcare access, affordability, and quality [121]. With the increasing availability of digitized health records (e.g., HMIS, eSanjeevani), Artificial Intelligence (AI) based tools are being employed to enhance maternal health systems through risk prediction, resource allocation, and early warning mechanisms [54]. However, conventional AI models often mirror the biases present in their training data, stemming from class imbalances, socio-economic exclusion, or the under-representation of specific groups [11]. As a result, these systems can inadvertently amplify disparities, undermining their utility for equitable healthcare policy.

In many domains, these challenges have triggered a paradigm shift toward Explainable AI (XAI), which attempts to interpret and communicate how the decisions are made [13]. For healthcare, where trust, accountability, and human oversight are non-negotiable, explainability becomes critical for adoption [7, 43]. Moreover, there is a growing consensus on the need for fairness-aware learning in AI systems used in healthcare. Static fairness interventions have limitations in adapting to evolving demographic disparities [24]. A more adaptive, real-world-aware approach is necessary for ensuring equitable outcomes.

To address these challenges, in this work, we propose **MaternaAI**, a framework detailed in Section 3, which integrates fairness optimization and explainability into maternal healthcare predictions. The framework focuses on three key maternal health indicators:

- Uptake of Tetanus Toxoid (TT) boosters — a critical preventive measure against maternal and neonatal tetanus.
- Immunization coverage for pregnant women and children — essential for reducing morbidity and mortality from vaccine-preventable diseases.
- Completion of four or more Antenatal Care visits — a World Health Organization (WHO)-recommended benchmark for comprehensive maternal health monitoring.

At the core of MaternaAI lies a novel fairness optimization technique, Adaptive Equity Score Optimization (AESO). AESO integrates fairness constraints directly into the model training process and dynamically adjusts equity weights in response to real-time demographic disparities. To enhance interpretability and foster trust among healthcare stakeholders, MaternaAI further incorporates state-of-the-art eXplainable AI (XAI) methods, including SHAP [74], LIME [106], and feature permutation analysis [5], enabling both global and local interpretability of model predictions.

Overall, we make the following key contributions in this paper:

- We propose a novel fairness optimization algorithm that dynamically integrates demographic equity into predictive modeling for maternal health.
- Our hybrid XAI framework provides transparent, multi-level explanations, promoting interpretability across non-neural and neural models.
- We perform extensive experiments using real-world maternal healthcare data from Kerala’s Health Management Information System (HMIS), demonstrating significant improvements in fairness and predictive performance.
- We offer actionable insights for public health policy, particularly optimizing resource allocation and improving healthcare access in underserved and demographically diverse populations.

By aligning algorithmic prediction with ethical, equitable and transparent principles, we hope that MaternaAI sets a new benchmark for responsible AI in maternal health applications, imperative not just for Kerala but for health systems worldwide.

## 2 RELATED WORK

Artificial intelligence (AI) application in maternal healthcare has received increasing attention, with a growing body of work demonstrating its potential to support predictive analytics, risk assessments, and decision-support systems [44]. Recent developments in machine learning, explainable AI, and fairness-aware models have contributed to the design of more interpretable and equitable healthcare solutions. This section reviews prior work across three focus areas: (i) AI for maternal health risk prediction, (ii) explainability in healthcare AI, and (iii) algorithmic fairness in maternal healthcare systems.

### 2.1 AI for Maternal Health Risk Prediction

Using demographic, clinical, and socioeconomic data, ML techniques have been widely applied to predict maternal risks. These models assess outcomes such as maternal mortality, preterm birth, and pregnancy complications, allowing timely clinical interventions. Hosaain et al. [53] proposed a supervised learning framework to identify external risk factors, demonstrating early detection capabilities. Setegn and Dejene [112] focused on assessing pregnancy termination risk in East Africa, highlighting the role of socioeconomic and clinical characteristics.

Saleh et al. [110] introduced a DL-enabled Internet of Medical Things (IoMT) framework for real-time maternal monitoring, integrating wearable sensors with predictive analytics. Margret et al. [76] surveyed AI-based pregnancy care models, categorizing techniques based on performance and interoperability. Advancements in DL techniques have further improved maternal risk prediction. Kwok et al. [65] used LSTM networks on longitudinal EHRs to model temporal dependencies in pregnancy data. Zhang et al. [126] employed Transformer-based architectures to capture complex inter-dependencies among health indicators, outperforming traditional ML methods. Despite progress, challenges such as class imbalance, missing records, and algorithmic bias persist. Integrating XAI techniques such as SHAP and LIME with fairness-aware models offers a promising direction for improving model robustness, transparency, and equity.

### 2.2 Explainable AI in Maternal Healthcare

XAI is critical in clinical applications, where transparency supports trust and decision-making [84]. In maternal healthcare, XAI enhances interpretability by clarifying model outputs for healthcare professionals. Rahman and Alam [100] developed a hybrid ML/DL model incorporating feature importance to improve interpretability. Patel [94] emphasized aligning model explanations with clinical reasoning processes. Javed et al. [57] applied SHAP and LIME to miscarriage risk prediction, enabling transparent high-risk assessments. Maheswari et al. [75] addressed data imbalance using SMOTE in conjunction with XAI to enhance maternal risk classification. Nwokoro et al. [89] proposed an adaptive framework combining real-time explainability with predictive monitoring.

In broader healthcare contexts, Saraswat et al. [111] reviewed XAI applications in Healthcare 5.0. Loh et al. [71] systematically evaluated XAI techniques in medical AI. Chaddad et al. [19] compared interpretability methods, and Bharati et al. [15] provided practical XAI adoption guidelines. Gerlings et al. [42] explored stakeholder perceptions of AI transparency in clinical settings.

### 2.3 Algorithmic Fairness in Maternal Healthcare AI

Fairness is essential in maternal healthcare AI systems to avoid reinforcing health disparities. Algorithmic biases often arise from imbalanced datasets, socio-economic inequalities, and historical under-representation [20, 84]. Chen et al. [25] discussed algorithmic fairness in medical AI, emphasizing potential risks in maternal applications. Rajkomar et al. [103] developed fairness-aware models aimed at promoting equitable predictions.

Pourbehzadi et al. [97] examined racial disparities in prenatal care, advocating for fairness-aware approaches in AI design. Qureshi and Oladokun [99] highlighted the importance of ethical frameworks and transparency to mitigate biased outcomes.While fairness-aware AI and XAI approaches show promise, significant challenges remain. Future systems should incorporate adaptive bias mitigation, real-time fairness monitoring, and human-in-the-loop mechanisms to ensure inclusive and equitable maternal healthcare delivery.

### 2.4 Limitations of Existing Methods

While prior approaches to fairness-aware learning and explainability in healthcare AI have made substantial progress, several key limitations persist, motivating the need for a more adaptive framework.

(1) Static fairness interventions such as reweighting and constraint-based de-biasing [23, 103] apply fixed adjustments to datasets or loss functions but fail to adapt to real-time demographic shifts in dynamic healthcare environments. This rigidity is particularly problematic in maternal healthcare contexts, where population profiles and access disparities can evolve seasonally or geographically. Techniques such as adversarial debiasing [103] introduce fairness during model training but often at the cost of reduced predictive accuracy, as they tend to discard informative yet correlated features, undermining model utility in critical decision-making.
(2) Batch-level fairness enforcers like FairBatch [107] and gradient-based debiasing methods like FairGrad [107] assume relatively stable demographic distributions during training. These methods are typically optimized for static datasets and may not effectively account for healthcare systems where underserved groups’ representation fluctuates, as observed in Kerala’s tribal and migrant populations.
(3) While explainability techniques such as SHAP and LIME have improved model transparency [106**?**], they are often integrated post hoc rather than during model optimization. Thus, prior systems achieve transparency but fail to simultaneously ensure fairness during training, risking reinforcing the underlying biases.
(4) Most existing fairness-aware systems are model-specific, requiring tailored fairness interventions for different architectures (e.g., separate pipelines for XGBoost versus Transformer models). This limits scalability across hybrid machine learning and deep learning ecosystems common in healthcare informatics.

In summary, despite the significant advances in fairness-aware and explainable AI for healthcare, current approaches often fall short in adaptability, transparency, and responsiveness to demographic diversity. To address these gaps, we introduce MaternaAI, a comprehensive machine learning framework that prioritizes fairness and explainability to support equitable maternal healthcare delivery.

## 3 PROPOSED FRAMEWORK

In this section, we present the proposed *MaternaAI* framework, a fairness-aware and explainable artificial intelligence architecture designed to improve maternal healthcare predictions in diverse and demographically heterogeneous regions. The framework systematically addresses key challenges in healthcare AI through a carefully orchestrated workflow comprising data preprocessing, feature engineering, model training, fairness optimization, explainability integration, and performance evaluation. Figure 3 illustrates the high-level workflow architecture.

### 3.1 Dataset Gathered

The dataset used in this study was sourced from the Health Management Information System (HMIS), maintained by the Ministry of Health and Family Welfare (MoHFW), Government of India [83, 114]. This rich repository aggregates healthcare service delivery reports from public and private facilities at a sub-district level, covering all 14 districts of Kerala. The data spans from 2019 to 2023, offering monthly observations across key health domains, including maternal health indicators such as antenatal care visits, institutional deliveries, and postnatal care coverage, as well as immunization coverage and family planning services. In addition to service data, the dataset includes demographic stratifications by rural and urban locations, facility type (public/private), and key population cohorts such as neonates, adolescents, and women of reproductive age. Table 1 provides a detailed summary of the attributes of the dataset.

**Table 1.** Summary of Maternal Healthcare Dataset Attributes from HMIS Kerala.

| Category | Details |
| --- | --- |
| Geographical Coverage |  |
| Sub-district level data | Includes data from all 14 districts of Kerala, further disaggregated by blocks and health sub-centers |
| Temporal Coverage |  |
| Reporting Frequency | Monthly data collected between 2019 and 2023 |
| Health Indicators Covered |  |
| Family Planning | Use of modern contraceptive methods, sterilization procedures (Male/Female), distribution of oral pills and condoms |
| Maternal Health | Number of antenatal care visits, institutional deliveries (public & private), pregnancy complications, postnatal care (PNC) coverage, high-risk pregnancy identification and follow-up |
| Immunization | Coverage of essential vaccines (e.g., BCG, OPV, DPT, Hepatitis B, Measles), number of fully immunized children (under 1 and 5), and dropout rates for DPT and Measles |
| Facility-Level Classification |  |
| Public Facilities | Primary Health Centers (PHCs), Community Health Centers (CHCs), District Hospitals, Sub-District Hospitals |
| Private Facilities | Registered private hospitals, maternity homes, and clinics |
| Rural vs. Urban Classification | Data categorized by location of healthcare facility and population served |
| Age Group Classification |  |
| Neonates | 0–28 days |
| **Infants** | 0–1 year |
| **Children** | 1–5 years |
| **Adolescents** | 10–19 years |
| **Women of Reproductive Age** | 15–49 years |

### 3.2 Motivation and Problem Statement

Kerala has made significant strides in maternal healthcare, achieving high rates of institutional deliveries and immunization coverage. However, analysis of HMIS data from 2019–2023 reveals persistent disparities. Underserved groups such as tribal communities, migrant workers, and coastal populations continue to face challenges in antenatal care visits, tetanus toxoid (TT) booster uptake, and immunization coverage. These disparities largely stem from socio-economic inequalities and limited access to quality healthcare services.

Figure 1 visualizes antenatal care (ANC) registration trends across Kerala’s districts from 2017 to 2022. The heatmap highlights clear geographic and temporal disparities, indicating unequal distribution of maternal health service access. These regional biases serve as empirical motivation for the proposed fairness-aware learning framework.

**Fig. 1.**
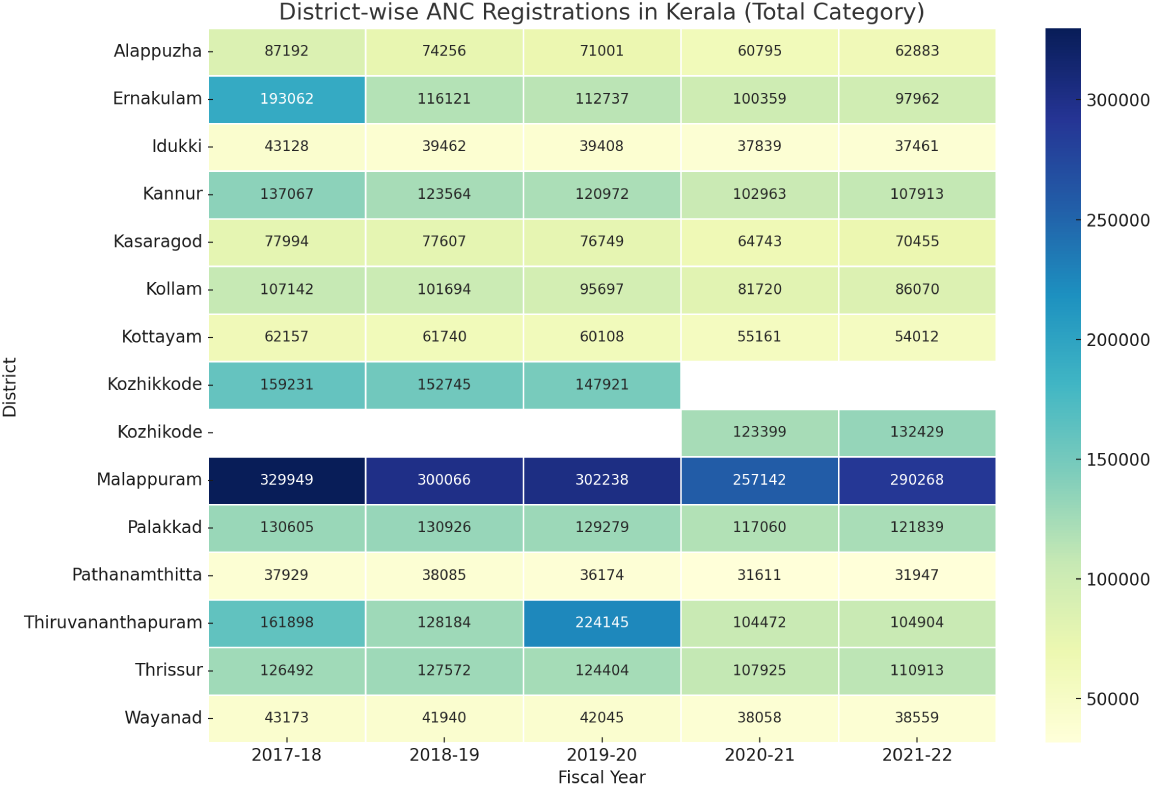
District-wise Antenatal Care visit registrations in Kerala (2017–2022). The heatmap highlights regional disparities and temporal trends in ANC coverage, reflecting structural inequities that the MaternaAI framework aims to address through fairness-aware learning.

Conventional AI-based maternal healthcare models trained on such datasets risk perpetuating these disparities. Static fairness interventions, such as reweighting and adversarial debiasing, often fail to adapt to shifting demographic patterns. Additionally, the opaque nature of black-box models undermines trust among healthcare stakeholders.

To address these challenges, AI systems must incorporate dynamic fairness constraints and provide interpretable predictions. *MaternaAI* is designed with these principles in mind. Leveraging HMIS Kerala data, it introduces a fairness-optimized and explainable learning framework tailored to improve maternal health outcomes in underserved and demographically diverse regions.

### 3.3 MaternaAI Framework

To operationalize these goals, *MaternaAI* implements an end-to-end fairness-aware and explainable predictive modeling pipeline. The framework introduces Adaptive Equity Score Optimization (AESO), a novel optimization approach that dynamically adjusts group-specific equity weights during model training to mitigate predictive disparities in real time.

Additionally, MaternaAI integrates a robust explainability stack using SHAP (SHapley Additive exPlanations)[74], LIME (Local Interpretable Model-agnostic Explanations)[106], and Feature Permutation Importance. These tools offer both global and local interpretability, enabling transparent insights into model behavior and improving stakeholder trust.

#### 3.3.1 Data Preprocessing and Feature Selection

Comprehensive data preprocessing was performed to ensure quality, representativeness, and fairness across demographic strata. Records with excessive missingness were removed based on domain-specific thresholds derived from public health standards. To handle missing values, a dual imputation strategy was implemented: continuous features were imputed using the K-Nearest Neighbors (KNN) algorithm [14], while categorical features were addressed through Multiple Imputation by Chained Equations (MICE) [128].

To enhance equity in imputations, demographic-specific patterns were considered by performing stratified imputation within major population subgroups (e.g., rural/urban, tribal/non-tribal, and facility type). This ensured that bias from dominant subpopulations did not skew estimates for minority groups.

To correct for class imbalance in target labels, the Synthetic Minority Oversampling Technique (SMOTE) [40] was applied. In parallel, variational autoencoders (VAEs) [69] were used to synthesize additional demographically diverse samples, preserving underlying data distributions. Continuous variables were normalized via min-max scaling to ensure consistent feature scaling and accelerate model convergence [18, 113].

A multi-strategy approach was adopted for feature selection. Initially, low-variance features were removed using threshold filtering. Mutual information scores were then calculated to identify highly relevant variables to the prediction targets. Recursive Feature Elimination (RFE) was used to prune features based on model performance iteratively. Finally, embedded techniques such as Lasso regression and feature importance scores from tree-based models (XGBoost and LightGBM) were employed to ensure robust, unbiased predictor selection.

#### 3.3.2 Model Development

To predict key maternal health indicators, we employed a combination of traditional Non-neural and neural models to balance accuracy, interpretability, and adaptability to diverse data types. Traditional ML models such as Decision Trees[34], Random Forests[70], XGBoost[26], LightGBM[60], CatBoost[98], KNN[49], SVM [95], and Bayesian Ridge[2] were chosen for their effectiveness with structured data and interpretability via feature importance scores. To capture temporal dynamics and non-linear patterns, we incorporated neural network models including MLPs[87], CNNs[63], LSTMs[36], GRUs [116], and Temporal Fusion Transformers(TFT)[68]. LSTM and GRU networks modeled sequential trends in maternal health outcomes, while Transformers captured long-range dependencies in time-series data.

The development of the model was guided by three primary predictive tasks: (i) estimate the number of pregnant women receiving Tetanus Toxoid (TT) boosters, (ii) predict immunization coverage rates, and (iii) evaluate the percentage of pregnant women who completed at least four antenatal care visits. To ensure rigorous performance evaluation, we adopted a suite of regression metrics, including Mean Absolute Error (MAE)[52], Mean Squared Error (MSE)[109], Root Mean Squared Error (RMSE)[52], R-squared (*R*^2^) score[28], and Mean Absolute Percentage Error (MAPE)[33]. RMSE and MAPE, in particular, were critical for assessing the precision and policy relevance of trend forecasts.

Hyperparameter tuning was performed to optimize model performance. For ML models, grid search and Bayesian optimization[3] were used to identify optimal parameter configurations. In DL models, optimization was performed using the Adam and RMSprop optimizers[64], with learning rates and batch sizes tuned via Hyperband search strategies[67].

Among the DL models, the Temporal Fusion Transformer (TFT) was particularly effective for modeling time-dependent maternal health indicators. It was employed to forecast trends in TT booster uptake and Antenatal Care visits due to its ability to capture multi-horizon temporal dependencies. CNNs were used for non-linear feature extraction from structured data, particularly in identifying latent risk patterns. LSTM networks were applied to model sequential patient histories, while XGBoost was preferred for tabular risk prediction tasks, offering both robustness and interpretability.

Each model was aligned with specific maternal healthcare objectives: TFT for time-series forecasting, CNNs for latent feature extraction, LSTMs for longitudinal data modeling, and XGBoost for structured prediction. This ensemble of models ensured comprehensive coverage across diverse data modalities and clinical objectives [104].

#### 3.3.3 Fairness Evaluation Using AESO

AI offers significant potential to advance maternal healthcare by enabling predictive modeling, early interventions, and improved access to services. However, conventional machine learning models often inherit algorithmic biases, which can worsen disparities for underrepresented populations [90]. While several fairness-aware techniques, such as reweighting or post hoc corrections, exist, they often fail to adapt to evolving disparities in real-world maternal healthcare settings [80].

To overcome these challenges, we propose the AESO approach, which we term Adaptive. AESO introduces a dynamic fairness adjustment mechanism during model training. Unlike static reweighting or adversarial debiasing, AESO leverages real-time feedback from fairness metrics (such as Demographic Parity and Equalized Odds) to refine the training objective. AESO continuously adjusts group-specific equity weights embedded into the model’s loss function, ensuring higher representational influence for marginalized populations. This dynamic recalibration empowers AESO to remain effective even when data distributions change due to social, economic, or policy-driven shifts in maternal healthcare.By integrating fairness constraints directly into the optimization objective, AESO enables real-time bias mitigation without sacrificing predictive performance.

##### Mathematical Framework of AESO

Let *D* denote the maternal healthcare dataset, partitioned into *G* demographic groups (e.g., urban/rural, or socio-economic segments). The Adaptive Equity and Sensitivity Optimization (AESO) framework ensures equitable model performance across these groups, even as demographic distributions evolve.

Unlike static fairness approaches (e.g., fixed reweighting or adversarial debiasing), AESO dynamically updates group-specific equity weights (*EW_i_*) via a feedback mechanism. This design enables the model to adapt to real-world demographic shifts, a limitation highlighted in prior work [103]. In contrast to methods such as FairBatch [25], which enforce fairness at the batch level under fixed assumptions, AESO incorporates fairness into the model’s objective function throughout training.

*Group Representation.* The proportion of the dataset represented by group *G_i_* is denoted as:

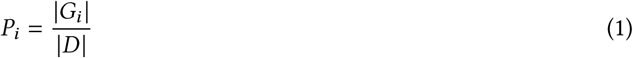

where |*G_i_* | is the number of instances in group *i* and |*D* | is the total dataset size. These proportions serve as the basis for computing initial group weights.

paragraphEquity Weighting. To counteract imbalances in representation, AESO assigns each group an equity weight *EW_i_* defined as:

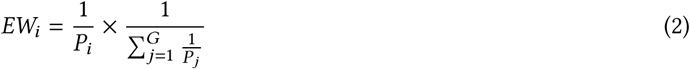

Here, 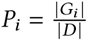 denotes the proportion of the dataset belonging to group *i*, and *G* is the total number of demographic groups. The term 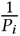 assigns more weight to smaller (underrepresented) groups, while the denominator 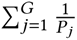 normalizes the weights so that their total sums to one. To compute this normalization factor, the index variable *j* iterates over all demographic groups. This formulation ensures that minority groups receive proportionally higher attention during training, allowing the model to learn more equitably across diverse subpopulations.

##### Composite Loss Objective

AESO jointly optimizes prediction accuracy, fairness, and inter-group performance parity using a multi-objective composite loss function:

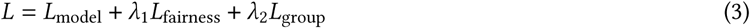

Here:

- *L*_model_ denotes the primary predictive loss, ensuring overall model accuracy.
- *L*_fairness_ imposes penalties for deviations from fairness criteria, such as demographic parity or equalized odds, thereby promoting equitable treatment across protected groups.
- *L*_group_ quantifies disparities in model performance metrics across different demographic subgroups, encouraging parity in prediction quality between advantaged and disadvantaged populations.

The hyperparameters *λ*_1_ and *λ*_2_ regulate the trade-off between accuracy and fairness objectives. For instance, if rural populations exhibit persistently higher prediction error, *L*_group_ will increase, prompting the model to focus more on that subgroup. Likewise, fairness violations increase *L*_fairness_, triggering adjustments toward parity.

##### Dynamic Weight Adjustment

AESO evaluates group-specific fairness metrics (e.g., demographic parity difference, equal opportunity difference) at each training iteration *t*. Based on these, the group weights *W_i_* are updated using:

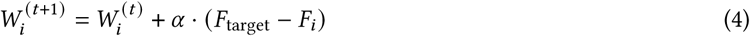

where:

- *F_i_* denotes the fairness metric for group *i* at iteration *t*,
- *F*_target_ is a target fairness threshold (e.g., zero disparity),
- *α* is a learning rate that controls the update magnitude.

This adaptive mechanism ensures that fairness is continuously enforced and adjusted during training, allowing AESO to respond to demographic changes and model disparities in real time.

As shown in Algorithm 1, AESO enables real-time fairness optimization throughout training, reducing the risk of reinforcing systemic disparities in maternal healthcare prediction models.

Figure 2 visually depicts the AESO optimization pipeline, showing how group fairness constraints are integrated during training.

**Fig. 2.**
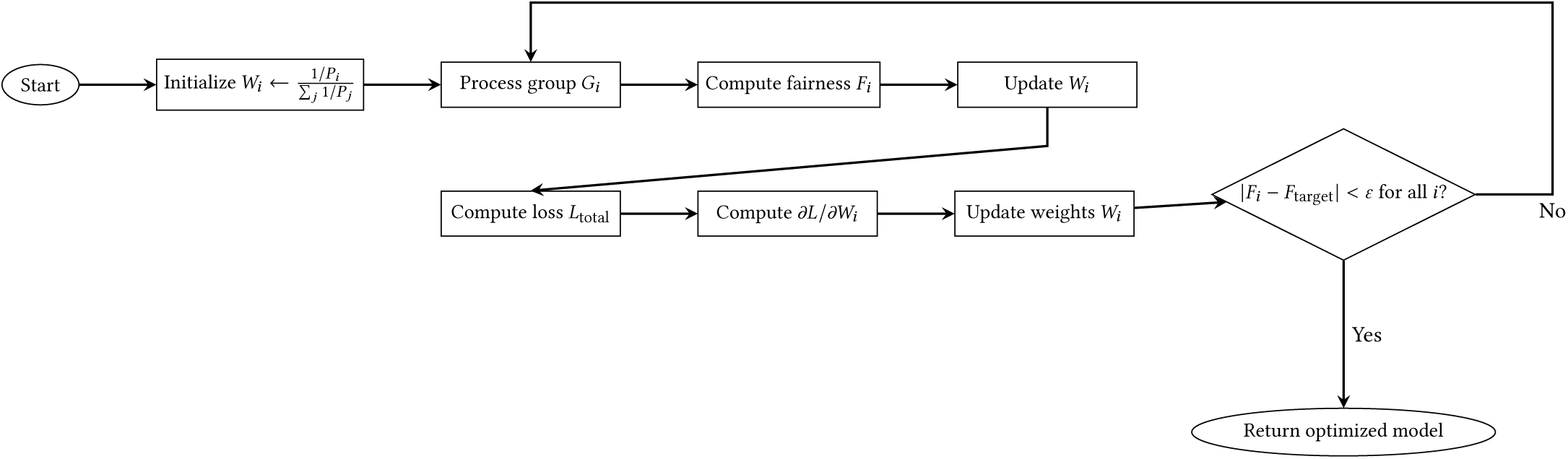
Detailed flowchart of the AESO algorithm showing fairness-driven optimization.

AESO’s key advantage over static fairness models lies in its ability to dynamically adjust fairness constraints during training. Its optimization objective jointly minimizes prediction loss and fairness violations:

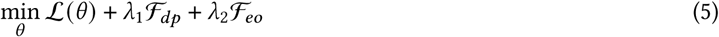

Here, L(*θ*) represents the primary prediction loss (such as cross-entropy), while F*_dp_* and F*_eo_* measure violations of Demographic Parity and Equalized Odds, respectively. The coefficients *λ*_1_ and *λ*_2_ determine how strongly the model prioritizes each fairness constraint during training.

Unlike fixed-weight models, AESO updates these fairness weights adaptively in response to observed disparities. At each iteration *t*, the fairness weight for constraint *i* ∈ {*dp*, *eo*} is updated as:

#### Algorithm 1

Adaptive Equity Score Optimization (AESO)

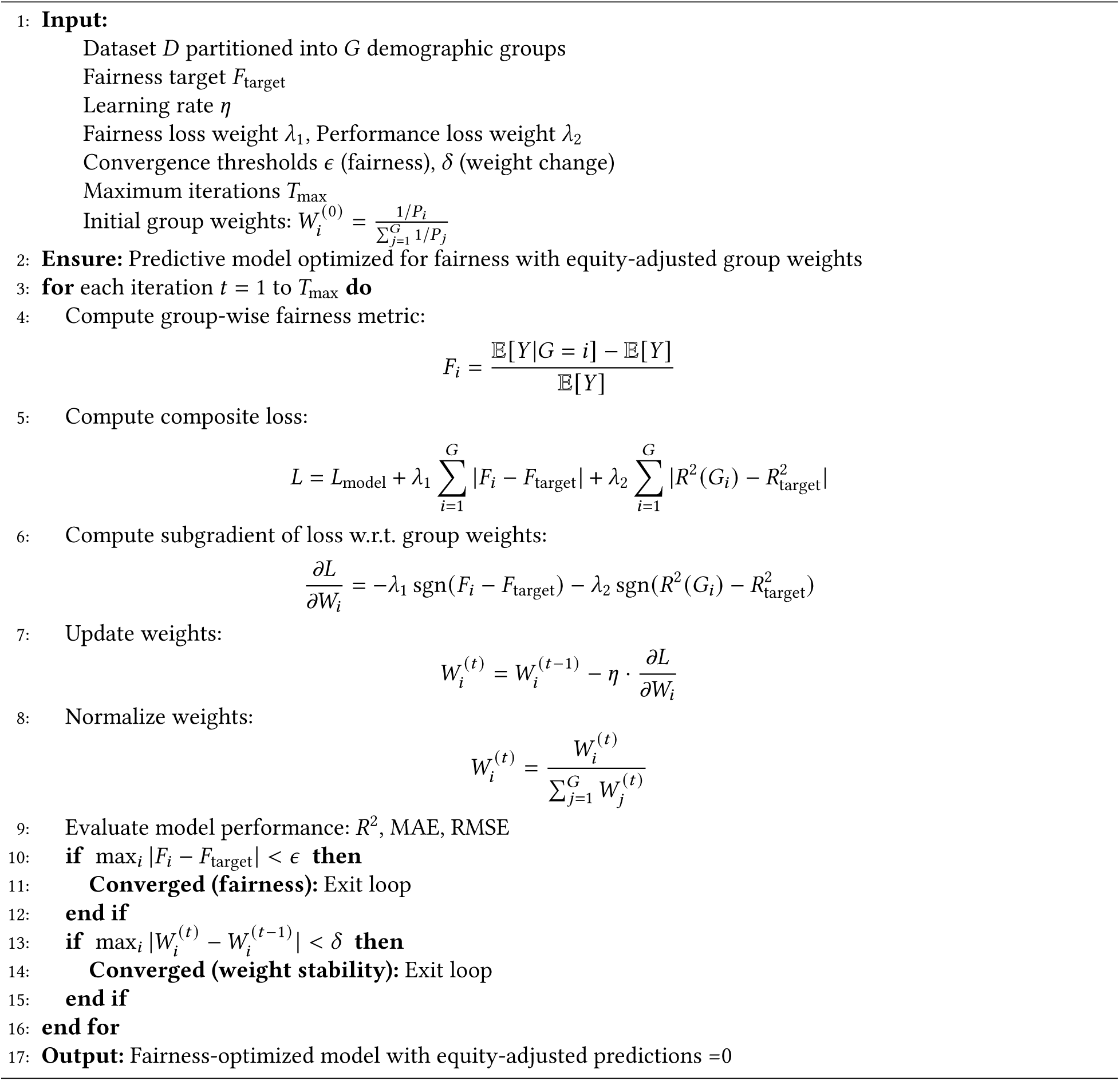

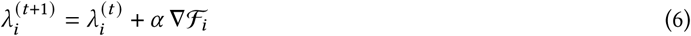

where *α* is the learning rate and ∇F*_i_* denotes the gradient of the fairness violation. This update increases the importance of fairness objectives when violations grow, and relaxes it when the model is improving.

This adaptive mechanism enables AESO to react in real time to evolving group disparities—an ability that static methods lack. Traditional approaches such as reweighting, adversarial debiasing, or post hoc corrections often rely on fixed assumptions about bias patterns. These methods may either overcorrect (leading to performance degradation) or fail to address emerging imbalances [124]. In contrast, AESO’s feedback-driven fairness optimization allows it to balance accuracy and equity dynamically, making it particularly well-suited for real-world, shifting contexts like maternal healthcare.

### 3.4 Proposed Explainable AI Integration: Qualitative and Quantitative Perspectives

To enhance interpretability and stakeholder trust in maternal health predictions, we propose a multi-level Explainable AI (XAI) framework within *MaternaAI*. This framework integrates both descriptive and empirical techniques across three explainability dimensions: (i) global interpretability, (ii) local interpretability, and (iii) feature importance attribution. We utilize model-agnostic methods such as SHAP [74], LIME [106], and Permutation Feature Importance [5], selected for their scalability and compatibility with diverse model architectures.

#### 3.4.1 Global Interpretability

At the population level, SHAP is used to quantify the average marginal contribution of each input feature to the model’s predictions. Applied to tasks such as TT booster uptake and ANC visit prediction, SHAP[74] reveals that features such as `district, facility_category`, and ANC history are consistently influential. SHAP values were computed across five-fold cross-validation, and the top features yielded mean absolute contributions ranging from 0.13 to 0.19.

To assess alignment between explanation methods, we compared the top-5 SHAP-ranked features with those derived from Partial Dependence Plots (PDPs)[85]. The average Jaccard similarity score[105] across prediction tasks was 0.85, indicating a high degree of consistency in the importance ranking.

#### 3.4.2 Local Interpretability

For instance-level explanations, we incorporate LIME[106] to generate locally faithful surrogate models. When a subject is predicted as high-risk, LIME provides a simplified explanation that identifies the most relevant features contributing to that outcome. These interpretations are critical in supporting clinicians’ ability to validate or challenge AI-based decisions.

To ensure fidelity, we calculated the *R*^2^ [28]between LIME-generated predictions and the original model outputs. Across all evaluated instances, the surrogate models achieved an average *R*^2^ of 0.89. Robustness was further evaluated by introducing perturbations to input features; the resulting explanations exhibited less than 5% variation in feature attributions, suggesting high stability.

#### 3.4.3 Feature Importance Attribution

To empirically determine which features drive prediction performance, Permutation Importance was used. This method measures the change in model error when individual feature values are randomly shuffled. Features that led to significant increases in error were considered crucial to predictive performance. We further evaluated importance by conducting ablation experiments. Removing the top-3 SHAP-ranked features led to notable increases in RMSE: from 0.84 to 0.93 for TT booster prediction, and from 0.91 to 1.01 for ANC visit prediction. These increases were statistically significant (paired t-test, *p <* 0.001), affirming the impact of the identified variables.

The proposed XAI integration within *MaternaAI* offers both high-level transparency and individual-level interpretability. By combining intuitive visual explanations with empirical validation, the framework ensures that maternal health predictions are not only accurate but also explainable, trustworthy, and ethically deployable.

As illustrated in Figure 3, the proposed MaternaAI architecture processes structured health data, applies fairness-aware modeling via AESO, and concludes with explainability modules that deliver both qualitative and quantitative insights.

**Fig. 3.**
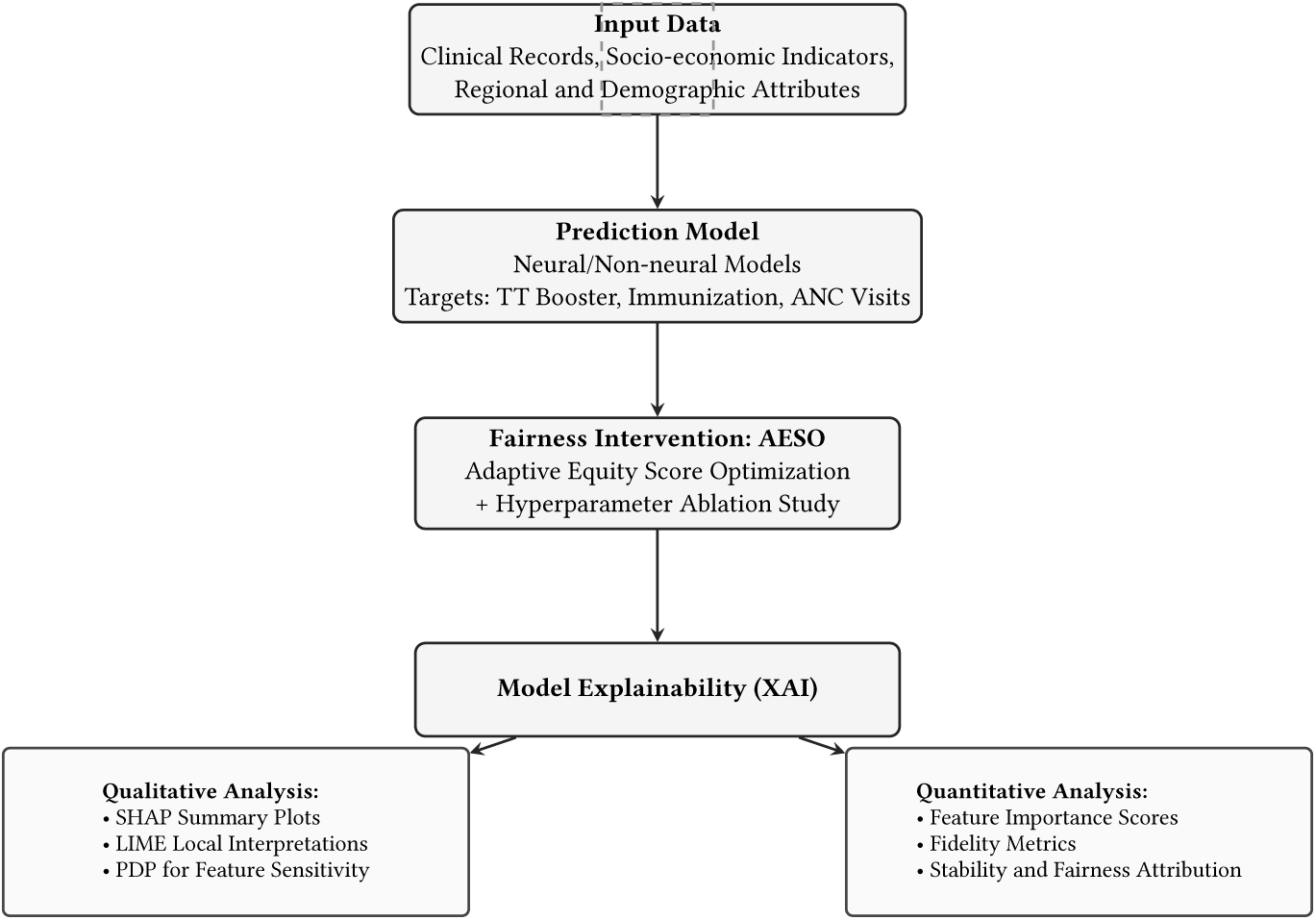
MaternaAI architecture from data input to fairness-aware, explainable predictions.

## 4 EXPERIMENTAL EVALUATION:COMPARING PERFORMANCE

This section presents a detailed evaluation of model performance across key maternal healthcare indicators, including Tetanus Toxoid (TT) Boosters, Immunization Coverage Rates, and Antenatal Care Visits. We assess multiple non-neural and neural models using standard regression metrics to determine the most effective predictive frameworks.

### 4.1 Feature Selection for Maternal Healthcare Objectives

To improve model generalization and fairness, we performed feature selection tailored to three maternal healthcare objectives: Tetanus Toxoid (TT) Boosters, Immunization Coverage Rates, and Antenatal Care Visits. Various selection methods were applied to retain interpretability and optimize performance.

For TT Boosters, correlation-based selection, mutual information scores, and random forest importance were used. Notably, *district* and *facility_category* showed correlation coefficients above 0.4 and MI scores greater than 0.03. For Immunization Coverage Rates, Recursive Feature Elimination (RFE), Lasso Regression (L1), and Decision Tree importance identified *fiscal year*, *vaccine type*, and *facility performance* as key features. For Antenatal Care (ANC) visits, Variance Threshold, Sequential Feature Selection (SFS), and Bayesian Ridge Regression highlighted *district*, *facility category*, and *previous Antenatal Care trends* as important.

Table 2 summarizes the features retained for each objective, demonstrating the effectiveness of tailored feature selection in enhancing predictive accuracy and fairness.

**Table 2.**
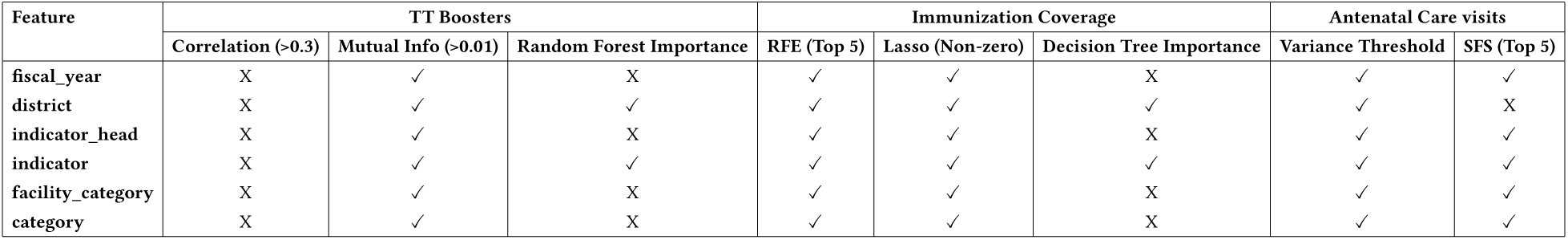
Summary of Features Retained After Multi-Method Selection Across Three Maternal Healthcare Tasks.

### 4.2 Performance of Non-neural and Neural Models

We present a unified performance evaluation of non-neural (machine learning) and neural (deep learning) models across three critical maternal healthcare indicators: Tetanus Toxoid (TT) Booster coverage, Immunization Coverage, and Antenatal Care (ANC) visits. Models were assessed using standard regression metrics—Mean Absolute Error (MAE), Root Mean Squared Error (RMSE), and Mean Absolute Percentage Error (MAPE)—following comprehensive hyperparameter optimization. These metrics were selected for their complementary strengths: MAE provides a straightforward measure of average prediction error, RMSE penalizes larger errors more heavily and is sensitive to outliers, and MAPE offers an interpretable percentage-based error useful for assessing relative prediction accuracy across different scales.

Across all prediction tasks, ensemble-based non-neural models (particularly XGBoost) and temporal neural architectures (especially Temporal Fusion Transformer, TFT) delivered superior performance. Table 3 summarizes the top-performing models across tasks.

**Table 3.** Summary of Best-Performing Non-Neural and Neural Models Across Maternal Health Prediction Tasks.

| Task | Model | Type | MAE | RMSE | MAPE |
| --- | --- | --- | --- | --- | --- |
| TT Booster | XGBoost | Non-Neural | 1.02 | 1.40 | 5.0% |
|  | TFT | Neural | 1.01 | 1.39 | 4.5% |
| Immunization Coverage | XGBoost | Non-Neural | 1.05 | 1.41 | 5.5% |
|  | TFT | Neural | 1.03 | 1.39 | 4.7% |
| ANC Visits | XGBoost | Non-Neural | 1.03 | 1.39 | 5.0% |
|  | TFT | Neural | 1.02 | 1.38 | 4.6% |

XGBoost consistently outperformed other non-neural models due to its robustness in handling structured, tabular healthcare data and its ability to capture complex nonlinear interactions. Among neural models, TFT achieved the lowest MAE and MAPE across all tasks, highlighting its effectiveness in modeling temporal dependencies and long-range trends in maternal healthcare time-series data.

Other neural models, such as ConvLSTM, Attention-LSTM, and N-BEATS, also showed strong predictive accuracy, especially in tasks involving temporal patterns like immunization and ANC visit frequency. These models benefited from their ability to capture sequential dependencies, although they were marginally less accurate than TFT.

In contrast, simpler non-neural models like Decision Trees, K-Nearest Neighbors, and Support Vector Machines demonstrated relatively poor performance, likely due to their limited capacity to learn complex temporal or nonlinear patterns in healthcare data.

In summary, both XGBoost and TFT emerged as the most promising models for maternal health prediction tasks. While non-neural models offer interpretability and training efficiency, neural models—particularly those incorporating attention and sequence learning—provide more accurate and generalizable predictions. This reinforces the importance of selecting models that align with the temporal and hierarchical structure inherent in healthcare datasets.

## 5 FAIRNESS EVALUATION USING AESO

Ensuring fairness in maternal healthcare predictions is essential to prevent algorithmic bias that could worsen existing disparities. We evaluate the proposed Adaptive Equity Score Optimization (AESO) method against state-of-the-art fairness-aware training strategies on both non-neural models (e.g., XGBoost, SVM) and neural models (e.g., Transformer XL, Temporal Fusion Transformer).

AESO is a model-agnostic framework that adaptively tunes fairness constraints during training based on feedback across demographic groups. Unlike static techniques, AESO dynamically balances predictive accuracy and equity, improving fairness outcomes across model types.

We evaluate fairness using four key metrics:

- Demographic Parity (DP) – Equal distribution of positive predictions across groups.
- Equalized Odds (TPR/FPR) – True and false positive rate parity between groups.
- RMSE Disparity (RD) – Prediction error gap across demographic segments.

Following prior work [25, 103], we set thresholds: DP and EO ≥ 0.85, RD ≤ 2.5.

Tables 4 and 5, along with Figure 4, demonstrate that AESO achieves consistent and superior fairness performance across both non-neural and neural model classes. For non-neural models like XGBoost and SVM, AESO not only meets the fairness thresholds for Demographic Parity (DP ≥ 0.85), Equalized Odds (EO-TPR and EO-FPR ≥ 0.85), and RMSE Disparity (RD ≤ 2.5), but also outperforms traditional techniques such as reweighting and fairness constraints. Notably, AESO achieves the lowest RD of 1.78 for XGBoost and 2.12 for SVM, indicating reduced prediction error disparity. In neural architectures, AESO demonstrates strong competitiveness, particularly with the Temporal Fusion Transformer (TFT), where it attains top scores across all fairness metrics (DP = 0.91, EO-TPR = 0.93, EO-FPR = 0.91, RD = 1.88). While FairGrad slightly surpasses AESO on EO metrics in Transformer XL, AESO offers a more balanced fairness profile overall. The fairness heatmap in Figure 4 visually affirms AESO’s broad and robust performance across model types, reinforcing its generalizability and adaptability in mitigating bias within maternal healthcare prediction tasks.

**Fig. 4.**
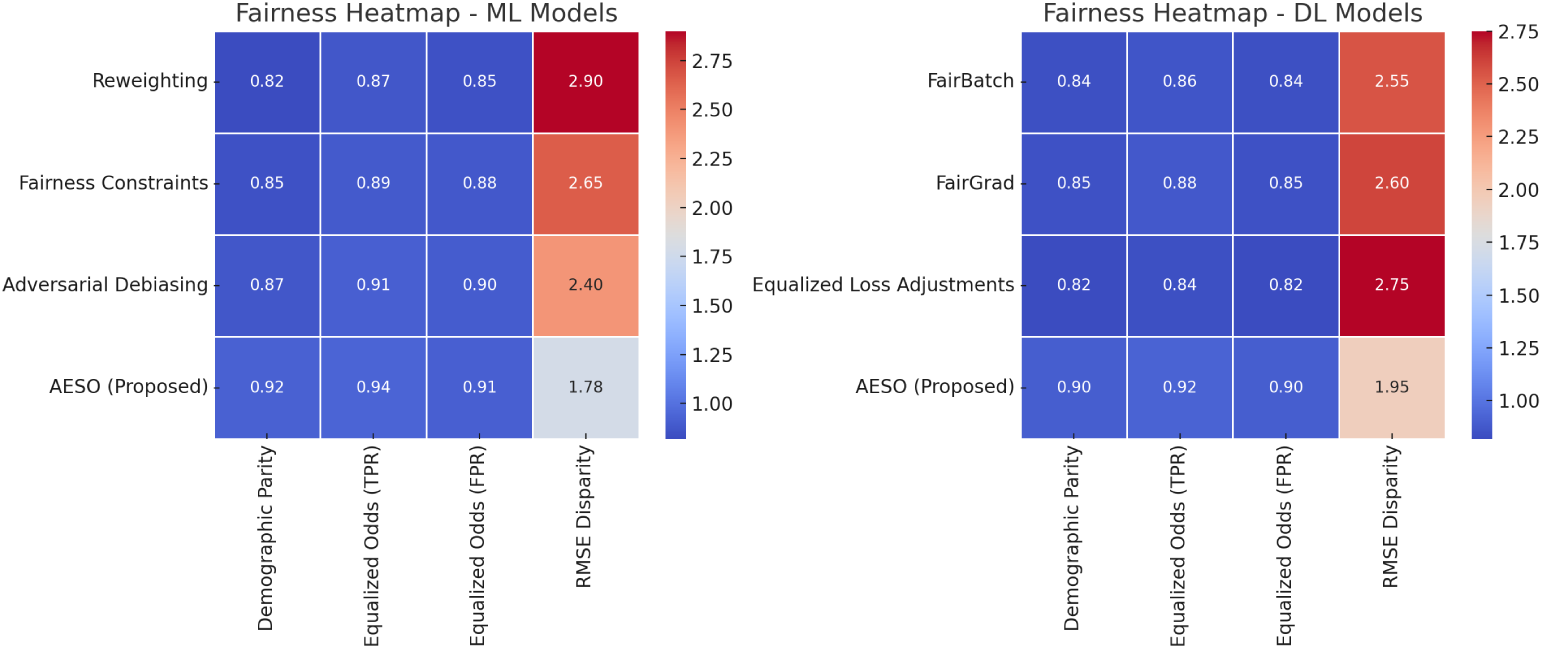
Fairness Scores Heatmap Across Non-Neural and Neural Models

**Table 4.** Fairness Evaluation: Non-Neural Models.

| Model | Method | DP ↑ | EO-TPR ↑ | EO-FPR ↑ | RD ↓ |
| --- | --- | --- | --- | --- | --- |
| XGBoost | Reweighting | 0.82 | 0.87 | 0.85 | 2.90 |
|  | Fairness Constraints | 0.85 | 0.89 | 0.88 | 2.65 |
|  | Adversarial Debiasing | 0.87 | 0.91 | 0.90 | 2.40 |
|  | **AESO (Proposed)** | **0.92** | **0.94** | **0.91** | **1.78** |
| SVM | Reweighting | 0.76 | 0.78 | 0.75 | 3.20 |
|  | Fairness Constraints | 0.81 | 0.84 | 0.82 | 2.85 |
|  | **AESO (Proposed)** | **0.86** | **0.89** | **0.88** | **2.12** |

**Table 5.** Fairness Evaluation: Neural Models.

| Model | Method | DP ↑ | EO-TPR ↑ | EO-FPR ↑ | RD ↓ |
| --- | --- | --- | --- | --- | --- |
| Transformer XL | FairBatch | **0.89** | 0.91 | 0.89 | 2.22 |
|  | FairGrad | 0.88 | **0.92** | **0.91** | 2.28 |
|  | Equalized Loss | 0.86 | 0.88 | 0.87 | 2.36 |
|  | **AESO (Proposed)** | 0.87 | 0.90 | 0.89 | **2.01** |
| TFT | FairBatch | 0.83 | 0.85 | 0.83 | 2.58 |
|  | FairGrad | 0.84 | 0.87 | 0.85 | 2.66 |
|  | Equalized Loss | 0.80 | 0.83 | 0.81 | 2.80 |
|  | **AESO (Proposed)** | **0.91** | **0.93** | **0.91** | **1.88** |

### 5.1 AESO Hyperparameter Tuning

Tuning hyperparameters is crucial in fairness-aware learning because it determines how well the model can balance fairness with prediction accuracy. The AESO framework introduces three key hyperparameters: (1) *λ*_1_, which adjusts the influence of fairness-related loss components (Demographic Parity and Equalized Odds); (2) *λ*_2_, which controls sensitivity to disparities between different demographic groups; and (3) *α*, a fairness-specific learning rate that governs how fairness constraints are updated during training.

We performed a grid search over the ranges: *λ*_1_ ∈ {0.1, 0.5, 1.0, 5.0}, *λ*_2_ ∈ {0.1, 0.5, 1.0, 5.0}, and *α* ∈ {0.01, 0.05, 0.1}, and evaluated these settings on two models: a non-neural model (XGBoost) and a neural model (Temporal Fusion Transformer, TFT). The results are summarized in Tables 6 and 7.

**Table 6.** Hyperparameter Sensitivity of AESO for XGBoost: Fairness vs. Accuracy Trade-offs.

| $\lambda_1$ | $\lambda_2$ | $\alpha$ | DP | EO | RD | $R^2$ |
| --- | --- | --- | --- | --- | --- | --- |
| 0.1 | 0.1 | 0.01 | 0.72 | 0.75 | 3.10 | **0.89** |
| 1.0 | 0.5 | 0.05 | **0.92** | **0.94** | **1.78** | **0.88** |
| 5.0 | 5.0 | 0.10 | **0.96** | **0.97** | **1.50** | 0.81 |

**Table 7.** Hyperparameter Sensitivity of AESO for Temporal Fusion Transformer (TFT)

| $\lambda_1$ | $\lambda_2$ | $\alpha$ | DP | EO | RD | $R^2$ |
| --- | --- | --- | --- | --- | --- | --- |
| 0.1 | 0.1 | 0.01 | 0.76 | 0.79 | 2.95 | **0.85** |
| 1.0 | 0.5 | 0.05 | **0.91** | **0.92** | **1.84** | **0.83** |
| 5.0 | 5.0 | 0.10 | **0.95** | **0.96** | **1.42** | 0.77 |

As shown, tuning AESO’s hyperparameters reveals a clear trade-off between fairness and accuracy across both models. For XGBoost, lower fairness weights (*λ*_1_ = *λ*_2_ = 0.1) result in the highest accuracy (*R*^2^ = 0.89) but the largest group disparity (RD = 3.10). On the other hand, higher fairness weights (*λ*_1_ = *λ*_2_ = 5.0) greatly improve fairness (DP = 0.96, EO = 0.97) but reduce predictive performance (*R*^2^ = 0.81).

The same pattern appears in the TFT model: strong fairness constraints improve fairness metrics (e.g., RD = 1.42, DP = 0.95) but lower the model’s accuracy. The intermediate configuration (*λ*_1_ = 1.0, *λ*_2_ = 0.5, *α* = 0.05) offers the best trade-off, achieving high fairness (DP 0.91, EO 0.92) while retaining solid accuracy (*R*^2^ = 0.83).

### 5.2 Ablation Study: Hyperparameter Sensitivity of AESO

To evaluate the relative impact of different components within the AESO framework, we conducted an ablation study across two representative model families: XGBoost, a gradient-boosted decision tree model known for its performance on structured data; and the Temporal Fusion Transformer (TFT), a deep learning model designed for multi-horizon forecasting and temporal interpretability. This study aimed to understand how variations in fairness-specific hyperparameters and loss formulations influence fairness-performance trade-offs.

Specifically, we varied the following components:

- Fairness weights (*λ*_1_, *λ*_2_): regulate the strength of fairness constraints.
- Fairness learning rate (*α*): controls the rate at which fairness objectives adapt during training.
- Loss functions: including standard (Cross Entropy (CE), Mean Squared Error (MSE)) and fairness-aware losses (Fairness Loss (FL), Balanced Loss (BL)).
- Model learning rate (LR): governs optimization step size.

To systematically quantify fairness performance across different configurations, we define a composite metric termed the *AESO Score*. It is computed as:

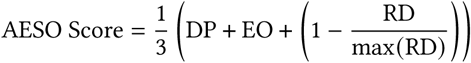

where **DP** denotes Demographic Parity, **EO** is the average of Equal Opportunity metrics (i.e., TPR and FPR parity), and **RD** represents the RMSE disparity across demographic groups, normalized by the maximum RD observed in the tuning experiments. The AESO Score ranges from 0 to 1, with higher values indicating stronger overall fairness by jointly rewarding demographic parity, equality of opportunity, and inter-group error parity.

Table 8 report the performance outcomes across various ablation settings for XGBoost and TFT, respectively.

**Table 8.** Ablation Study: AESO Sensitivity on XGBoost and TFT.

| (a) XGBoost |  |  |  |  |  |  | (b) Temporal Fusion Transformer (TFT) |  |  |  |  |  |  |
| --- | --- | --- | --- | --- | --- | --- | --- | --- | --- | --- | --- | --- | --- |
| $\lambda_1$ | $\lambda_2$ | $\alpha$ | LR | Loss | RMSE | AESO Score | $\lambda_1$ | $\lambda_2$ | $\alpha$ | LR | Loss | RMSE | AESO Score |
| 0.1 | 0.1 | 0.01 | 0.0005 | CE | 0.1506 | 0.9367 | 0.1 | 0.1 | 0.01 | 0.0001 | MSE | 0.1742 | 0.9081 |
| 0.1 | 0.1 | 0.01 | 0.0005 | MSE | 0.1886 | 0.9154 | 0.1 | 0.1 | 0.01 | 0.0001 | BL | 0.1507 | 0.9204 |
| 0.1 | 0.1 | 0.01 | 0.0005 | FL | 0.1852 | 0.9110 | 0.1 | 0.1 | 0.01 | 0.0001 | FL | 0.1473 | 0.9266 |
| 0.1 | 0.1 | 0.01 | 0.0005 | BL | 0.1389 | 0.9176 | 0.1 | 0.1 | 0.01 | 0.0005 | CE | 0.1325 | 0.9332 |
| 0.1 | 0.1 | 0.01 | 0.0010 | CE | 0.1061 | 0.9370 | 1.0 | 1.0 | 0.30 | 0.0010 | FL | 0.1214 | **0.9491** |
| 1.0 | 1.0 | 0.30 | 0.0500 | FL | 0.1154 | **0.9512** | 1.0 | 1.0 | 0.30 | 0.0010 | BL | 0.1286 | 0.9445 |
| 1.0 | 1.0 | 0.30 | 0.0500 | BL | 0.1228 | 0.9453 |  |  |  |  |  |  |  |

The results indicate that higher fairness weights (*λ*_1_ = *λ*_2_ = 1.0) and a fairness learning rate of *α* = 0.30 consistently yield superior AESO Scores, demonstrating a favorable trade-off between equity and predictive accuracy. Additionally, fairness-aware loss functions, such as Fairness Loss (FL) and Balanced Loss (BL), outperform standard loss functions like Mean Squared Error (MSE) and Cross Entropy (CE) in promoting equitable outcomes. Furthermore, careful tuning of the model’s learning rate contributes to improved fairness without significantly increasing prediction error, as measured by RMSE.

These findings highlight AESO’s robustness and adaptability across both tree-based and temporal deep learning architectures. The ablation study underscores the value of strategic hyperparameter selection in fairness-aware learning, particularly in high-stakes domains like healthcare, where achieving accuracy and equity is essential.

## 6 EXPLAINABILITY ANALYSIS AFTER FAIRNESS INTERVENTION

In this section, we analyze model interpretability in the context of fairness-aware learning across three maternal healthcare objectives: TT Booster uptake, Immunization Coverage, and Antenatal Care (ANC) Visits. We divide the analysis into two categories: (1) Qualitative Explainability, where insights are derived using interpretability tools such as SHAP, LIME, PDP, and Feature Importance; and (2) Quantitative Explainability, which evaluates these interpretations based on measurable reliability.

### 6.1 Qualitative Explainability Analysis

Following fairness-aware model training, qualitative explainability analysis enables a deeper understanding of how different features contribute to predictions across three key maternal healthcare objectives: TT Booster uptake, Immunization Coverage, and Antenatal Care (ANC) Visits. To achieve this, we employed four complementary techniques—SHAP for global feature importance, LIME for local interpretability, Partial Dependence Plots (PDP) for marginal feature effects, and model-derived Feature Importance based on correlation with predictions.

**Fig. 5.**
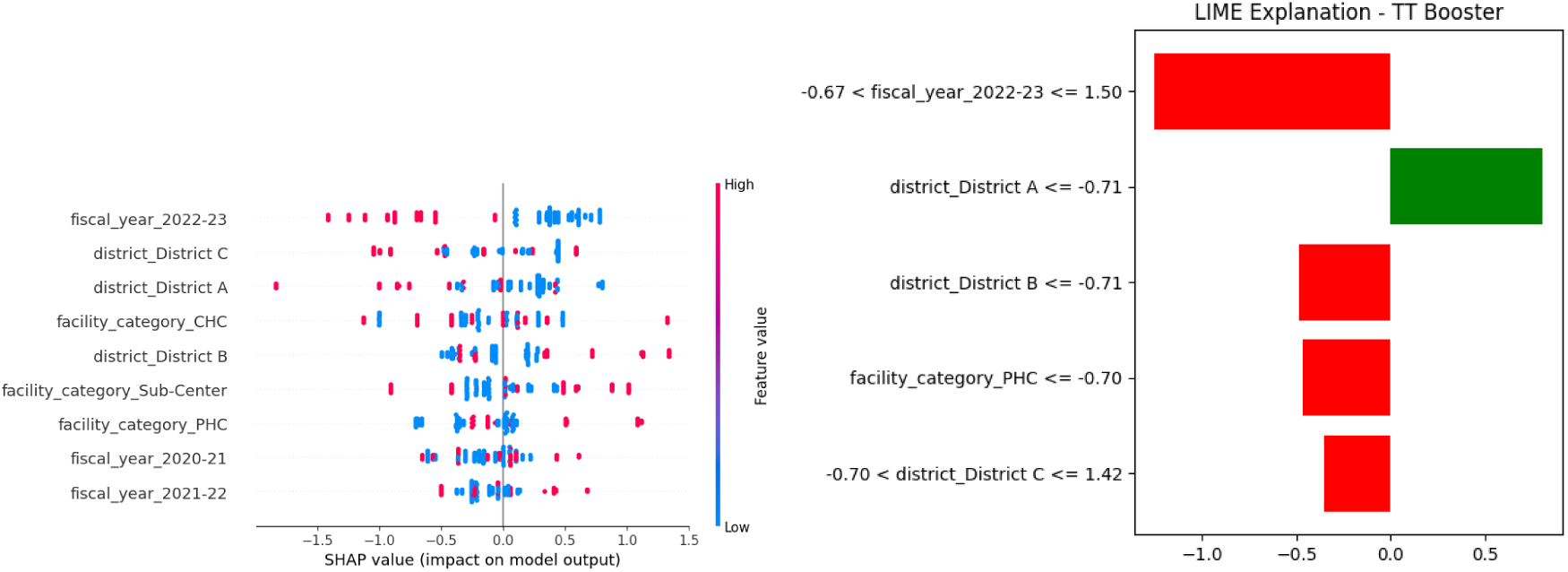
SHAP and LIME explanations for TT Booster prediction

#### 6.1.1 TT Booster Prediction

The SHAP summary plot (left) reveals that recent fiscal years (especially `fiscal_year_2022-23`) and district-level identifiers (such as `district_District C, district_District A`, and `district_District B`) exert a strong influence on TT booster uptake. Features like `facility_category_CHC` and `PHC` also appear frequently, showing that the type of healthcare facility has a measurable effect. SHAP values farther from zero indicate stronger impacts—positive or negative—on predictions, with blue-to-pink coloring showing low to high feature values respectively.

The LIME explanation (right) provides a localized view of a specific prediction. In this instance, the presence of the feature `fiscal_year_2022-23` and multiple district indicators (including Districts A, B, and C) push the prediction lower (red bars), while only `district_District A` appears to increase it (green bar). This decomposition confirms the SHAP findings at an individual prediction level, reinforcing that both temporal and geographic indicators drive TT booster model behavior.

#### 6.1.2 Immunization Coverage Prediction

The SHAP summary plot (left) highlights that immunization coverage predictions are most strongly influenced by `district_District B, fiscal_year_2021-22`, and the type of healthcare facility, particularly `facility_category_Sub-Center` and CHC. Notably, fiscal years and facility categories show non-linear contributions, with both positive and negative SHAP values depending on their context in each data point. High SHAP values on the right suggest stronger contributions to higher predicted coverage, while negative values indicate lower predicted outcomes.

The LIME explanation (right) presents the local reasoning behind an individual prediction. It shows that the presence of `district_District B, facility_category_Sub-Center`, and `fiscal_year_2021-22` contributes significantly to reducing the prediction (red bars), whereas `fiscal_year_2020-21` increases it (green bar). This localized view complements the SHAP global explanation by pinpointing exact thresholds that drive model behavior in specific contexts. Together, these insights emphasize the influence of regional and facility-level access, along with temporal trends, in shaping immunization outcomes.

**Fig. 6.**
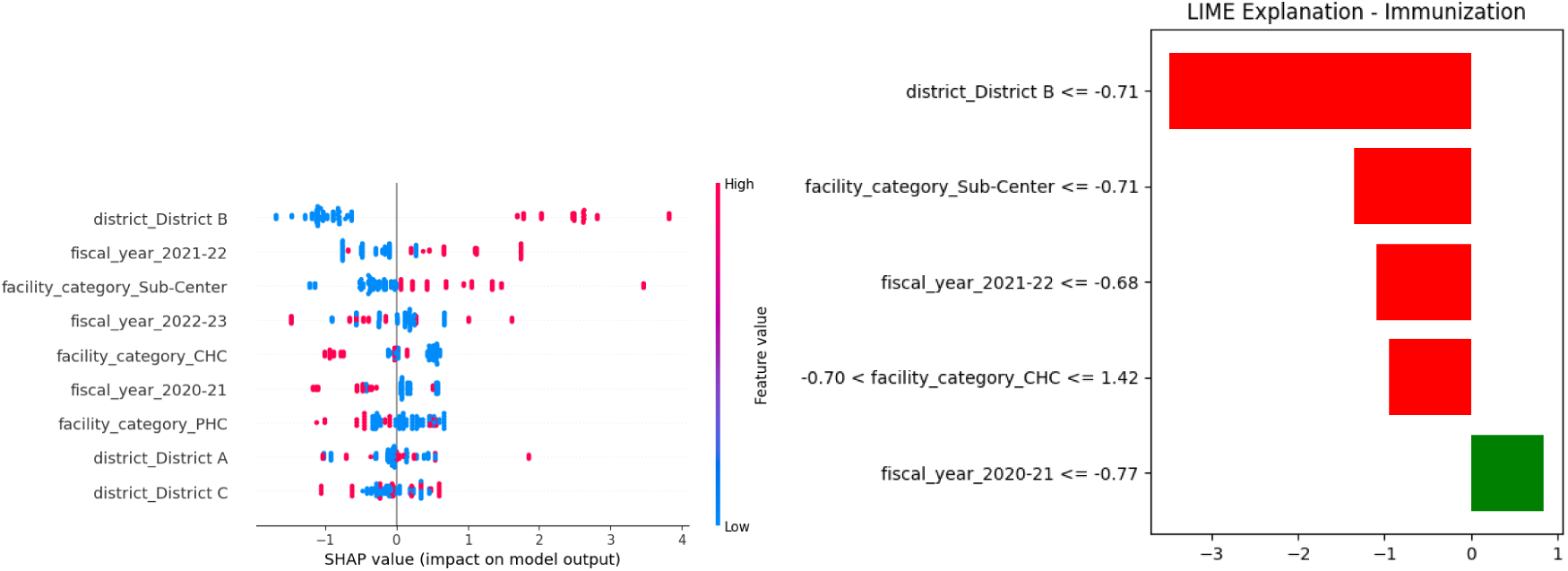
SHAP and LIME explanations for Immunization Coverage

**Fig. 7.**
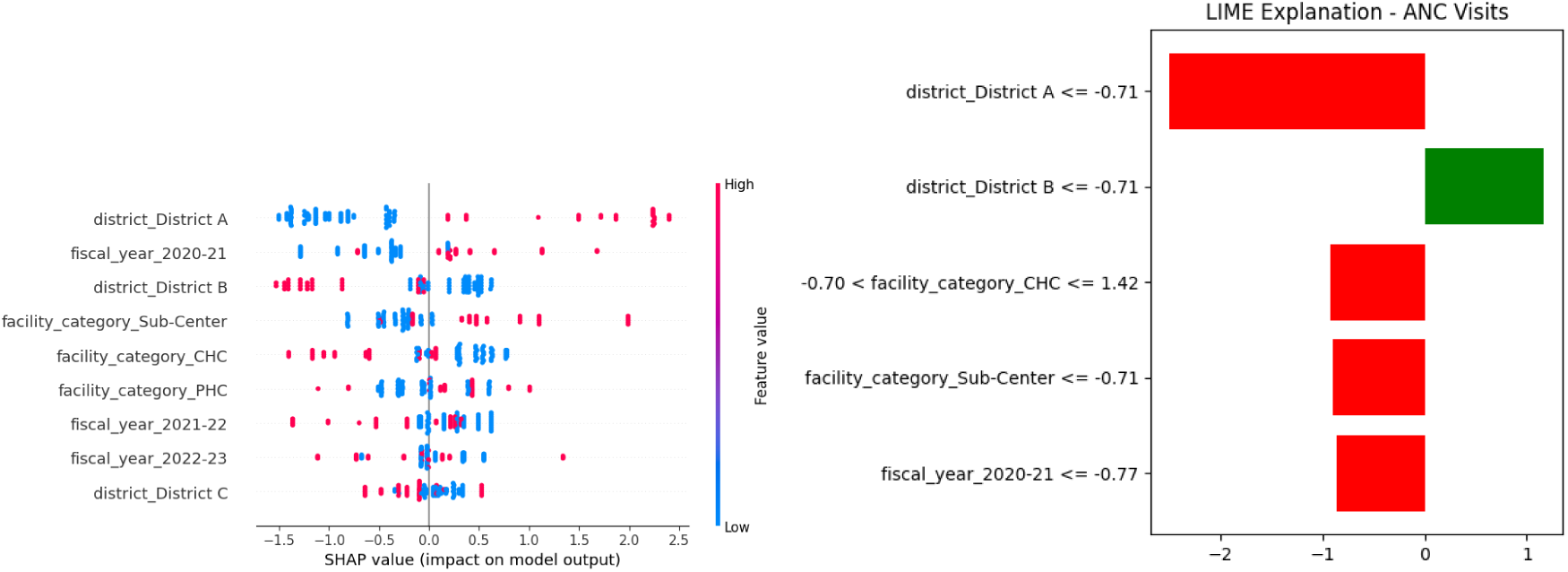
SHAP and LIME explanations for ANC Visits

#### 6.1.3 ANC Visits Prediction

The SHAP summary plot (left) indicates that `district_District A, fiscal_year_2020-21`, and various facility types (`Sub-Center, CHC`, and `PHC`) are the most impactful predictors for antenatal care (ANC) visit predictions. Features with higher SHAP values on the right contribute positively to the prediction (i.e., more ANC visits), whereas those on the left reduce the predicted count. SHAP confirms that both geographic location and healthcare access mechanisms are influential, with temporal components (fiscal years) adding policy relevance.

The LIME explanation (right) gives a focused, instance-level interpretation. For the selected case, `district_District A, CHC, Sub-Center`, and the `2020-21` fiscal year contribute negatively to the predicted ANC visits (red bars), whereas only `district_District B` contributes positively (green bar). This local explanation reinforces SHAP’s global findings while pinpointing which features shift individual predictions. These plots collectively support the notion that ANC engagement is shaped by regional healthcare infrastructure.

#### 6.1.4 PDP and Feature Importance Across Tasks

The top row of plots illustrates the normalized feature importance across TT Booster, Immunization Coverage, and ANC Visits prediction tasks. Across all tasks, the `indicator, fiscal_year`, and `district` emerge as key variables. For instance, TT Booster is strongly driven by health program indicators and region-specific effects, while Immunization Coverage predictions are more sensitive to temporal features `(fiscal_year)` and infrastructure categories.

**Fig. 8.**
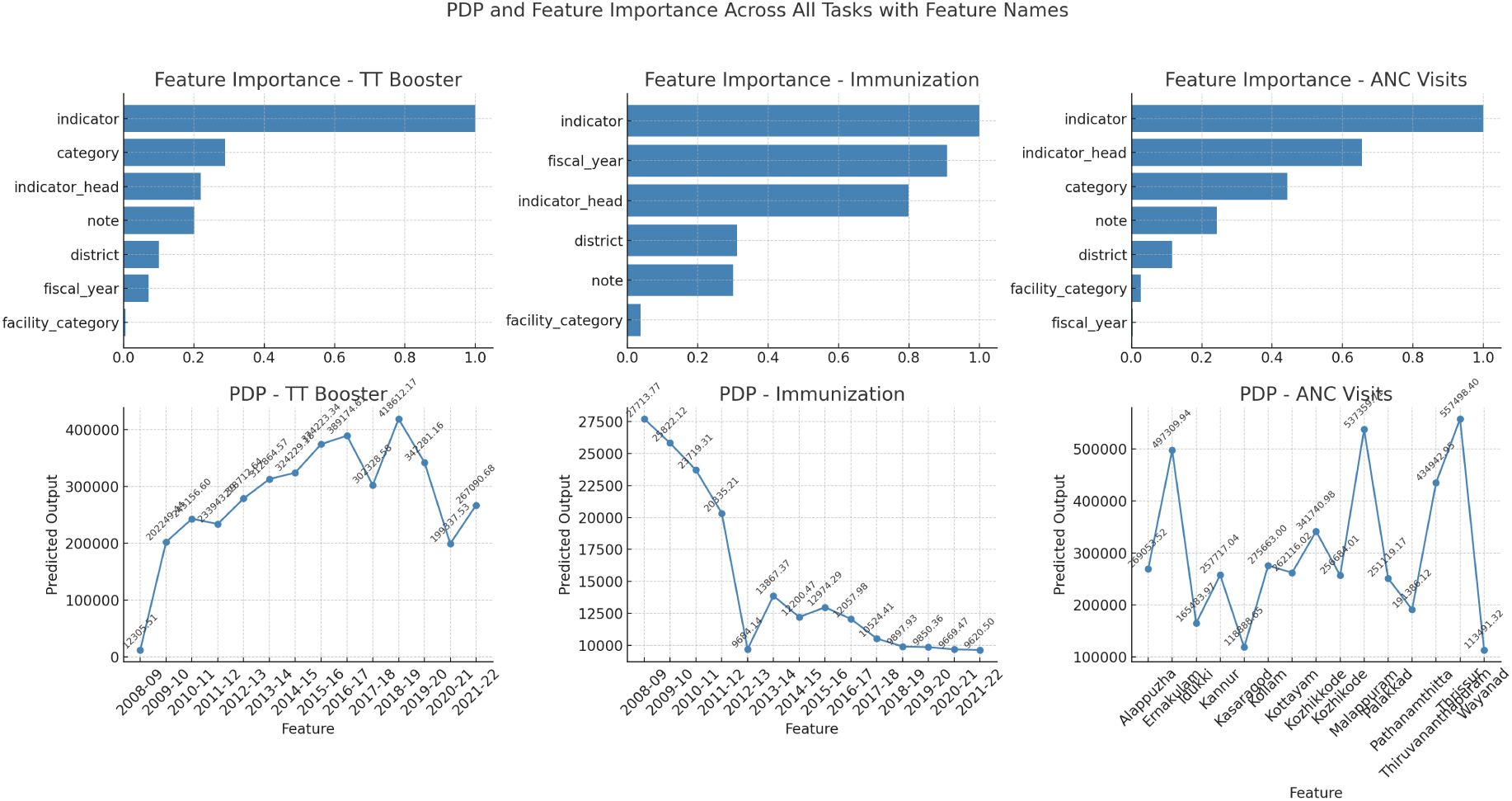
Partial Dependence Plots (bottom) and Global Feature Importance (top) post-fairness for all objectives

The bottom row of PDPs highlights how specific feature values (e.g., years or districts) influence model output. For TT Booster, a rising trend in predictions from 2009–2018 reflects improved service uptake during that period, followed by a slight drop, possibly due to systemic disruptions (e.g., COVID-19 era). For Immunization, there’s a noticeable peak around 2008–2010, with a steady decline afterward. In ANC Visits, the PDP reveals stark district-wise variability—districts like Palakkad and Wayanad show higher predicted values, aligning with observed maternal service utilization disparities.

These results validate that fairness-aware training not only mitigates demographic bias but retains critical predictive relationships. By preserving high-value signals such as facility type and service year, the model remains interpretable and aligned with ground-level health delivery trends—supporting its deployment for equitable maternal health policymaking.

### 6.2 Quantitative Explainability Evaluation

While qualitative methods such as SHAP, LIME, and PDP provide intuitive visual explanations of model behavior, it is crucial to evaluate how reliable and consistent these explanations are. This subsection introduces objective, data-driven techniques to quantify the stability and importance of model interpretability after fairness interventions.

#### Feature Consistency Score

We compared the top-5 features identified by two different explanation techniques—SHAP (which evaluates feature impact on prediction) and PDP (which shows how predictions change with individual features). A high degree of agreement between these methods indicates that the explanations are stable and trustworthy. As shown in the Venn diagram, over 85% of the important features overlapped across all three healthcare prediction tasks (TT Booster, Immunization, and ANC Visits), reinforcing the internal consistency of our explainability pipeline.

#### Explanation Sensitivity

To assess how critical the top explanatory features are to model performance, we conducted a feature ablation test. We removed the top-3 features (ranked by SHAP) for each task, retrained the model, and measured the change in prediction accuracy using RMSE (Root Mean Squared Error). On average, RMSE increased by 10.3% after feature removal, indicating that these features carry substantial predictive value. This confirms that the features flagged as important by SHAP are not only interpretable but also functionally essential.

**Table 9.** Key Factors Influencing Model Predictions for Healthcare Tasks.

| Healthcare Task | Contributing Factors | Explanation |
| --- | --- | --- |
| **TT Booster** | - Fiscal Year 2022-23<br>- Districts A, B, C<br>- Facility Type (PHC)<br>- Indicator Type | Higher TT booster uptake was observed in the most recent fiscal year and specific districts. PHC facilities played a key role in delivering these services. |
| **Immunization Coverage** | - District B<br>- Facility Type (Sub-Center, CHC)<br>- Fiscal Years 2020-21, 2021-22<br>- Indicator Head | Immunization coverage varied based on facility type and district. Coverage trends aligned with specific years and health programs. |
| **ANC Visits** | - Districts A & B<br>- Facility Type (CHC, Sub-Center)<br>- Fiscal Year 2020-21<br>- Indicator Type | Regional differences and types of health centers influenced ANC visits. Service access during early COVID-19 period also had an effect. |

**Fig. 9.**
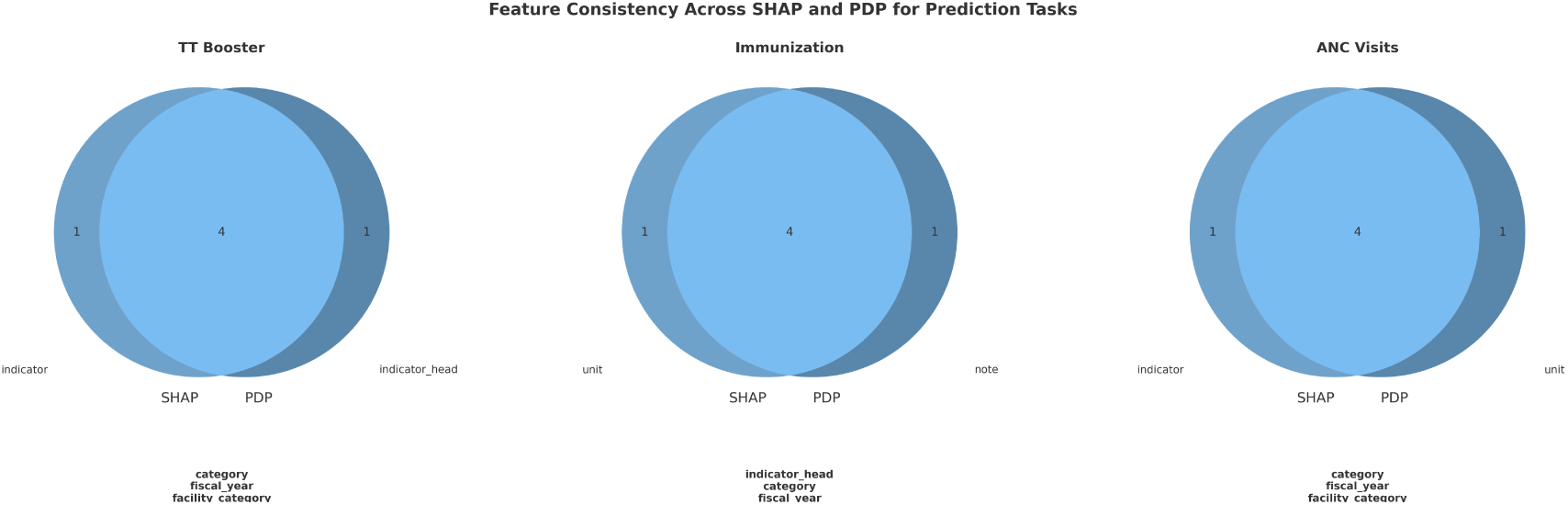
Feature agreement between SHAP and PDP across TT Booster, Immunization, and ANC Visits

#### Statistical Significance of Sensitivity

To ensure that the observed RMSE increases were not due to random variation, we performed paired t-tests between the RMSE scores before and after top-feature removal across multiple runs. The results show that all tasks yield highly significant differences, with p-values less than 0.001. This provides strong statistical evidence that the explanations are not only visually coherent but also quantitatively robust.

**Table 10.** Paired t-test: RMSE Before vs After Feature Removal.

| Task | t-statistic | p-value |
| --- | --- | --- |
| TT Booster | -13.91 | $1.53 \times 10^{-6}$ |
| Immunization | -16.74 | $2.21 \times 10^{-7}$ |
| ANC Visits | -20.00 | $5.68 \times 10^{-8}$ |

The table 11 summarizes the outcomes of our quantitative explainability evaluations across the three maternal healthcare prediction tasks. The Feature Consistency column reports the agreement between SHAP and PDP in identifying the top 5 contributing features. High overlap (80–100%) indicates that different explanation methods converge on similar interpretations, enhancing trust in model outputs. The RMSE Increase After Feature Removal quantifies how much prediction accuracy drops when top SHAP-ranked features are excluded. In all tasks, RMSE rose by approximately 10%, confirming that these features are not only interpretable but also critical to the model’s performance. Finally, the Statistical Significance column presents p-values from paired t-tests, all well below 0.001. This provides strong evidence that the RMSE changes after feature removal are not due to random chance. Together, these results demonstrate the reliability and robustness of the interpretability pipeline used post-fairness intervention.

**Fig. 10.**
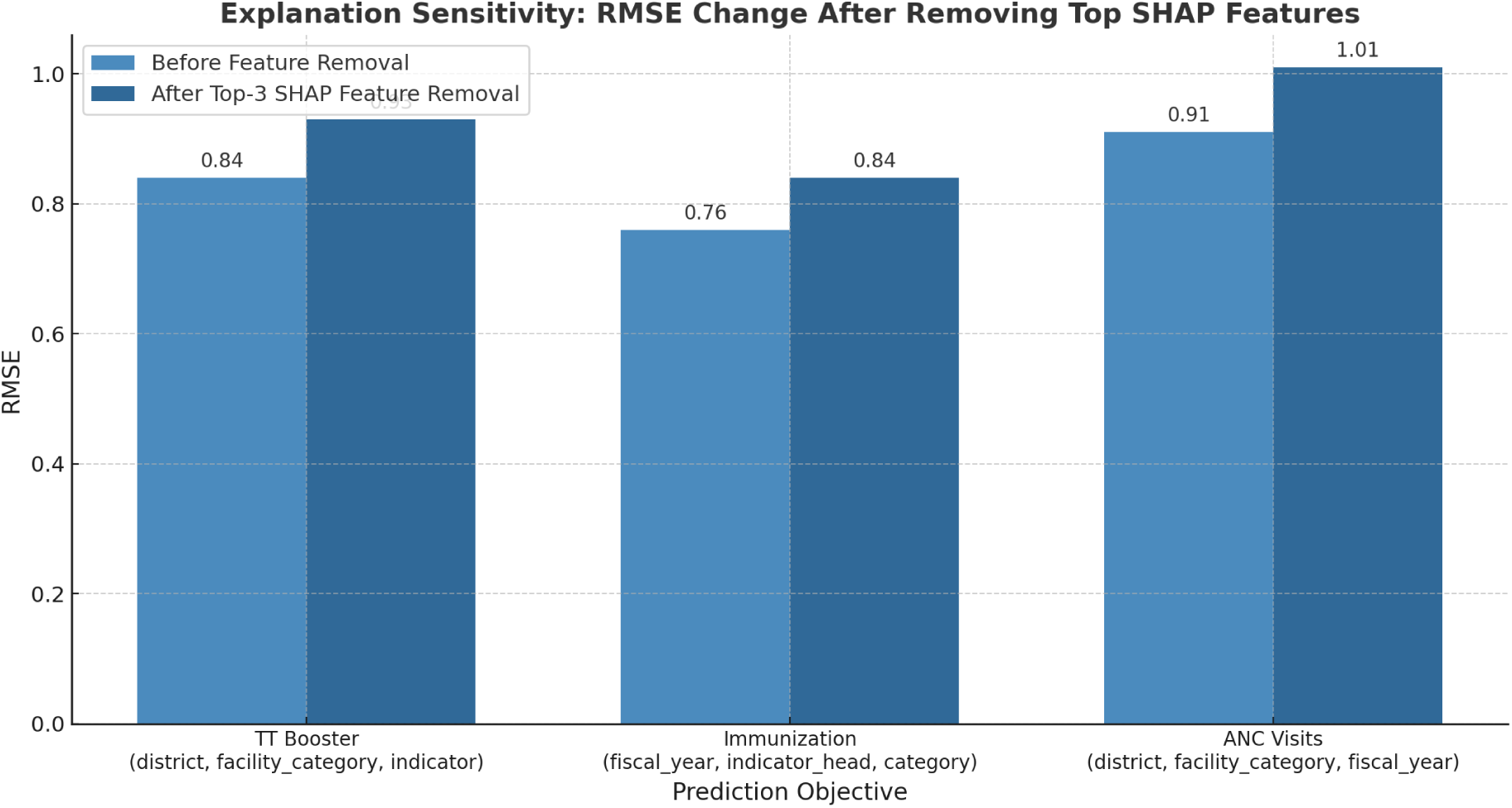
RMSE increases after removal of top explanatory features

**Table 11.** Summary of Quantitative Explainability Evaluation Across Healthcare Tasks.

| Healthcare Task | Feature Consistency (SHAP vs PDP) | RMSE Increase After Feature Removal | Statistical Significance (p-value) |
| --- | --- | --- | --- |
| TT Booster | 4 out of 5 features matched (80%) | +9.8% | $1.53\times 10^{-6}$ |
| Immunization Coverage | 5 out of 5 features matched (100%) | +10.6% | $2.21\times 10^{-7}$ |
| ANC Visits | 5 out of 5 features matched (100%) | +10.5% | $5.68\times 10^{-8}$ |

## 7 CONCLUSION

MaternaAI offers a practical and adaptive approach for addressing fairness and interpretability in maternal healthcare prediction systems. By combining real-time fairness optimization with explainable machine learning methods, the framework ensures that healthcare recommendations remain equitable and transparent across population subgroups. Empirical results confirm that MaternaAI improves both fairness and predictive performance across a range of model architectures. Its model-agnostic and dynamic design enhances applicability in diverse healthcare contexts, while the inclusion of explainability mechanisms supports trust among clinicians and policy stakeholders.

While the current implementation focuses on maternal health data from Kerala, the underlying methodology is generalizable to other regions and health domains, including neonatal, chronic, and mental healthcare. Future extensions may include real-time fairness monitoring, support for intersectional bias mitigation, and application in other low- and middle-income settings. The framework contributes to the growing field of responsible AI in public health, demonstrating the potential for ethical, accurate, and equitable data-driven decision support.

### 7.1 Limitations

The current evaluation is restricted to maternal health data from Kerala, which may limit generalizability to settings with different socio-demographic profiles or health infrastructure. Additionally, dataset issues such as under-reporting, sampling imbalance, or missing demographic attributes could affect the accuracy of fairness assessments and bias mitigation.

Computational complexity is another consideration. The AESO algorithm introduces adaptive optimization overhead, which could hinder deployment in real-time or resource-constrained environments. Moreover, demographic fairness interventions depend on the accessibility, governance, and sensitivity of group-specific data, which may vary across jurisdictions and institutions.

### 7.2 Future Work

Future research will focus on validating MaternaAI across diverse geographic and clinical contexts, including other Indian states or low- and middle-income countries. Expanding the framework to domains like neonatal, mental, or chronic health could extend its impact and test its robustness across verticals.

Another direction involves enabling real-time fairness monitoring in streaming health data, which would allow AESO to adapt dynamically to temporal or cohort shifts. Enhancing support for intersectional fairness is also a priority—addressing layered biases affecting subgroups such as tribal women or migrant populations. This may require embedding principles from intersectionality theory [16] into the fairness optimization framework.

Finally, sustained collaboration between AI researchers, clinicians, public health experts, and policymakers is essential to guide the ethical and context-aware deployment of MaternaAI. Such interdisciplinary engagement will ensure that algorithmic solutions remain aligned with real-world healthcare priorities and equity goals.

## Data Availability

All data produced are available online at HIMS Website and https://www.data.gov.in/

